# Uncertainty Aware Decision Support with Computationally Expensive Simulation Models: A Case Study of HIV Intervention Scenarios

**DOI:** 10.64898/2026.04.15.26350970

**Authors:** Arindam Fadikar, Anna Hotton, Pedro Nascimento de Lima, Raffaele Vardavas, Nicholson Collier, Kara Jia, Sara Rimer, Aditya Khanna, John Schneider, Jonathan Ozik

**Affiliations:** Decision and Infrastructure Sciences Division, Argonne National Laboratory, Lemont, IL, USA; Department of Medicine, University of Chicago, Chicago, IL, USA; Engineering & Applied Sciences, RAND, Arlington, VA, USA; CausalPaths Analytics LLC, Los Angeles, CA, USA; Department of Behavioral and Social Sciences, Brown University, Providence, RI, USA

## Abstract

Detailed agent-based simulations are increasingly used to support policy decisions, but their computational cost and complex uncertainty structure make systematic scenario analysis challenging. We present a data-driven, uncertainty-aware decision support (DDUADS) workflow for using stochastic simulation models as decision-support tools under limited computational budgets. The approach combines several established techniques—sensitivity screening, Bayesian calibration using simulation-based inference, and multi-surrogate model integration for translational efficiency—into a coherent pipeline that enables uncertainty-aware policy analysis. Rather than producing a single baseline, the calibration stage yields a posterior distribution over plausible model parameterizations, allowing flexible, uncertainty-aware forward projections. We demonstrate the DDUADS workflow on the INFORM-HIV agent-based model of HIV transmission in Chicago to evaluate potential disruptions in antiretroviral therapy (ART) and pre-exposure prophylaxis (PrEP) use. While the specific application is HIV modeling, the challenges and techniques described here arise in other simulation studies and can be applied to decision support in other domains.

## 1 INTRODUCTION

Detailed simulation models are increasingly used to inform policy decisions in complex systems where experimentation in the real world is infeasible or costly. In epidemiology, agent-based and network-based models are often used to study the spread of infectious diseases and evaluate the potential impact of interventions (Hooten and Wikle 2010; Ozik et al. 2021). These models can represent heterogeneous populations, behavioral dynamics, and interactions among individuals, enabling examination of counterfactual scenarios that would otherwise be difficult to observe. However, the same features that make these models useful for decision support also make them challenging to use in practice (Banks and Hooten 2021). High-dimensional parameter spaces, stochastic simulation outputs, and substantial computational cost complicate calibration, uncertainty quantification, and systematic evaluation of intervention strategies.

Once calibrated to observed data, such simulation models are typically used to project the effects of alternative policies or interventions, such as changes in treatment access or prevention strategies, on future outcomes. However, for these projections to be useful in robust decision-making (Lempert 2019), it is necessary to account for uncertainties from multiple sources, including uncertain parameters, stochastic simulator dynamics, and approximation methods, which poses substantial challenges (Robinson et al. 2013; Kimpton et al. 2025). Doing so is nontrivial due to both the computational burden of repeatedly evaluating expensive simulators and the absence of systematic frameworks for integrating uncertainty propagation across calibration and scenario analysis stages.

This paper develops a Data Driven Uncertainty Aware Decision Support (hereby, DDUADS) workflow to address these challenges when applying computationally intensive simulation models for decision analysis. The DDUADS workflow integrates several established and novel techniques that together enable systematic calibration, uncertainty propagation, and flexible and efficient scenario exploration. Specifically, the approach combines data-informed model specification (Naimi et al. 2017), global sensitivity screening for input dimension reduction (Morris 1991), Bayesian calibration using simulation-based inference (Dyer et al. 2024), and a novel, multi-surrogate approach to integrate model calibration with the exploration of intervention scenarios (Section 3.4.1). Although most of these components have been studied extensively in isolation, applying them together in large-scale simulation studies introduces practical difficulties related to computational cost, uncertainty management, and methodological integration. Our objective is to demonstrate how these components, when organized into a coherent pipeline that supports uncertainty-aware scenario analysis with complex stochastic simulators, provide practical decision support benefits that surpass their individual contributions.

We illustrate the DDUADS workflow using INFORM-HIV, an agent-based network model of HIV transmission in Chicago (Hotton et al. 2025). We present an example use-case in which we evaluated how changes in HIV care and use of biomedical prevention, operationalized as reductions in antiretroviral therapy (ART) and pre-exposure prophylaxis (PrEP) use, influence future HIV incidence and prevalence. Using the proposed workflow, we first translate observational data into appropriate model inputs and further reduce the dimensionality of the uncertain parameter space through global sensitivity screening. The model with the reduced parameter set is then calibrated to empirical estimates of HIV prevalence from surveillance data using simulation-based inference (SBI) techniques, producing a posterior distribution over parameter configurations consistent with observed prevalence. Finally, we use surrogate models to efficiently explore a range of ART and PrEP disruption scenarios while propagating uncertainty from the calibration stage.

Although the application considered here focuses on HIV transmission, the workflow addresses practical challenges that arise broadly when using detailed stochastic simulations for decision support. The combination of Bayesian calibration, uncertainty-aware projection, and surrogate-assisted scenario exploration while accounting for input uncertainty provides an adoptable framework for transforming computationally expensive simulation models into tools that can support policy analysis in epidemiology and other domains. The remainder of the paper is organized as follows. A brief overview of the INFORM-HIV model is provided in Section 2. The technical description of the DDUADS workflow components is provided in Section 3. Finally, concluding remarks are given in Section 4.

## 2 INFORM-HIV MODEL OVERVIEW

The Integrated Framework for Modeling Social Determinants of HIV Transmission (INFORM-HIV) is a stochastic, discrete-time agent-based network model designed to simulate HIV transmission among Black men who have sex with men and transgender women in Chicago. The model was developed to support public health decision making through counterfactual scenario analysis to understand how engagement in HIV care, use of biomedical prevention, and social determinants of health (SDOH) jointly influence HIV incidence and prevalence over time. INFORM-HIV combines several key components: a synthetic population representing the population of interest in terms of demographics and health states, dynamic sexual partnership networks, and mechanisms linking SDOH to behaviors and care engagement (Figure 1). For a detailed description of INFORM-HIV, see Hotton et al. (2025).

**Figure 1:**
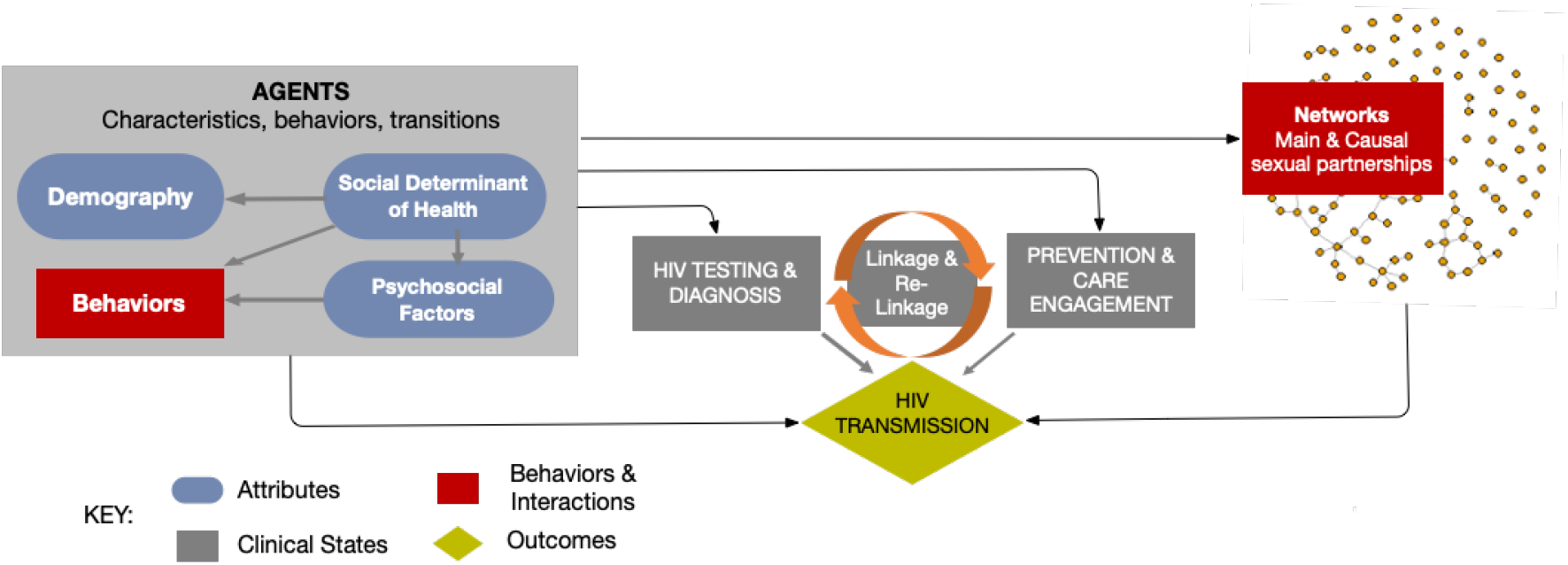
INFORM-HIV model schematics

The model represents individuals as agents with demographic attributes and health-related states relevant to HIV transmission and prevention. Each agent carries information such as HIV status, diagnosis status, engagement in care and viral suppression, and PrEP use. Additional attributes capture social determinants of health, including housing instability, incarceration, employment, and substance use. The simulator evolves in discrete daily time steps, updating agent states and partnership networks while recording epidemiological outcomes at specified intervals.

Sexual partnerships are represented as dynamic networks with main and casual relationships. Network formation and dissolution are simulated using separable temporal exponential random graph models (STERGMs), which allow the contact structure of the population to evolve over time while preserving realistic patterns of partnership formation. Conditional on partnership ties, HIV transmission events occur probabilistically as a function of per-act transmission risk, partnership type, contact frequency, and the infection status of each partner.

Several behavioral and biomedical factors modify transmission risk via changes in engagement in HIV prevention and care. ART use among HIV-positive agents can lead to viral suppression and reduced transmission probability, while PrEP use among HIV-negative agents reduces susceptibility to infection. Social determinants of health can influence these processes by affecting risk behaviors, partnership dynamics, and continuity of care. As a result, interventions in the model can operate through multiple pathways, including biomedical mechanisms such as ART and PrEP uptake as well as socio-structural changes that affect underlying SDOH conditions.

Key outputs from INFORM-HIV include time series of HIV prevalence and incidence, along with intermediate metrics that connect mechanisms to outcomes (e.g., diagnoses and care-cascade measures such as ART coverage and viral suppression, and PrEP uptake among HIV-negative individuals). The model can also generate stratified outcomes to support equity-relevant comparisons across subgroups and, if cost inputs are included, summaries relevant to economic evaluation.

## 3 DATA DRIVEN UNCERTAINTY AWARE DECISION SUPPORT (DDUADS) WORKFLOW

The DDUADS workflow provides a structured approach for using complex stochastic simulation models to support policy decisions under uncertainty by integrating observational data, statistical inference, and simulation-based modeling to produce predictive distributions under alternative interventions. Using INFORM-HIV as a motivating example and case study, we develop and describe each component of the workflow (shown in pink boxes in Figure 2), including the translation of observational data into model inputs, dimension reduction to identify influential parameters, Bayesian calibration to characterize posterior uncertainty, and surrogate-assisted scenario exploration for scalable what-if analysis. This framework is designed to address common challenges such as latent and weakly identifiable parameters and the computational burden of exploring high-dimensional intervention spaces, while maintaining a coherent probabilistic treatment of uncertainty throughout.

**Figure 2:**
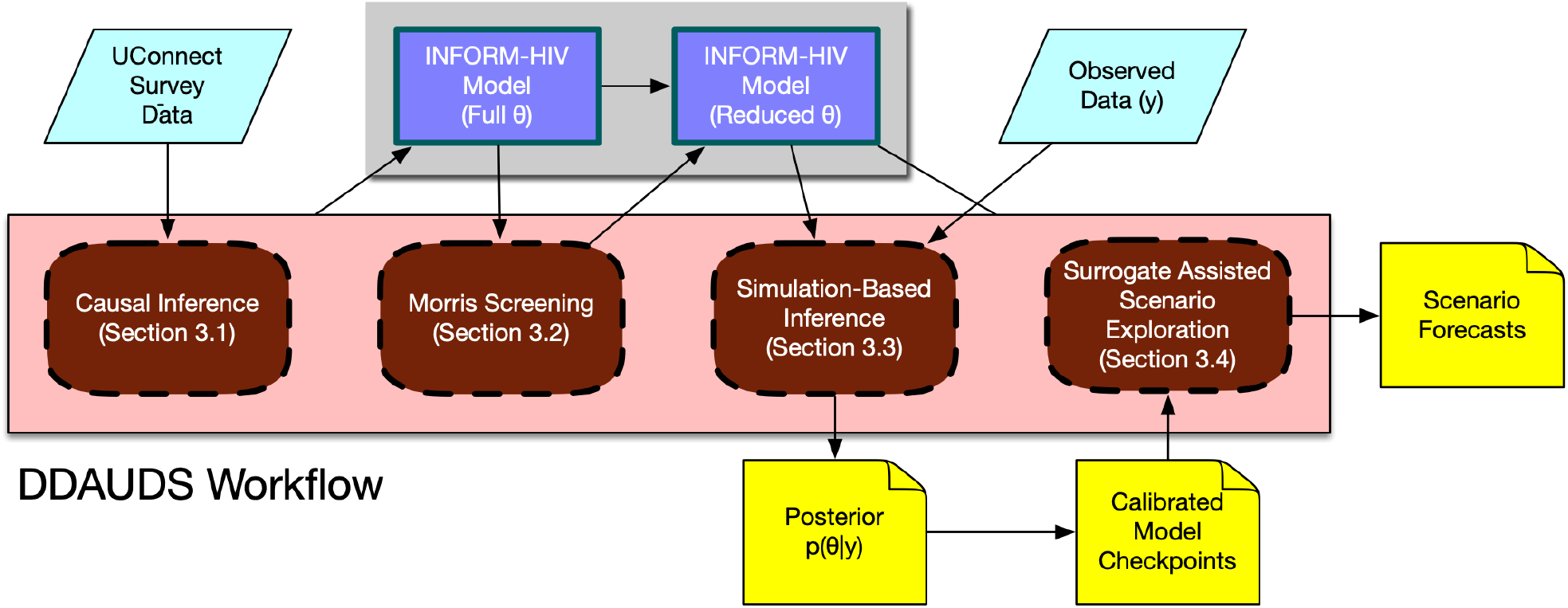
DDUADS Workflow connecting data-informed model construction, dimension reduction, calibration, and surrogate modeling for scenario analysis with INFORM-HIV. The boxes in brown represents the tools described in Section 3.

We now describe each component of the workflow in turn. In Section 3.1, we describe how observational data are translated into model inputs and model mechanisms. Next, Section 3.2 presents the global sensitivity screening procedure used to reduce the dimensionality of the parameter space and identify influential inputs. Section 3.3 then formulates and solves the Bayesian calibration problem to characterize the posterior distribution over parameters consistent with observed prevalence. Finally, Section 3.4 describes surrogate-assisted scenario exploration, which enables evaluation of intervention outcomes over a continuous decision space while propagating uncertainty from both calibration and emulation using an integrated, multi-surrogate model approach. We provide a companion repository that houses the software to enable each of the DDUADS workflow components (Fadikar 2026).

### 3.1 Connecting Survey Data to Simulator Mechanisms Via Causal Inference

Mechanistic simulation models are parameterized using a mix of (i) quantities that can be derived directly from available observations and (ii) quantities that cannot be measured directly and therefore must be estimated indirectly. In this application, an important modeling step concerns SDOH and how they are represented in the synthetic population. We use individual-level survey data that include HIV status and SDOH measures (e.g., housing instability, employment, substance use) to link observed information to two parts of the simulator. First, we use survey distributions and co-occurrence patterns to assign baseline SDOH states to agents in the initial population so that the synthetic population is consistent with observed joint structure. Second, we use the same data to determine how SDOH states evolve over time during the simulation, including dependencies where one SDOH factor affects the evolution of another (for example, housing affecting employment, or incarceration affecting housing).

Data used to parameterize the model came from a longitudinal study conducted in Chicago between 2013 and 2015 (the UConnect study) that examined the impact of social influences on HIV risk and use of biomedical and behavioral prevention among young Black men who have sex with men and transgender women (Schneider et al. 2017; Schneider et al. 2017).

The key challenge is that survey variables do not map one-to-one to simulator mechanisms. The model requires operational quantities such as transition probabilities, whereas the survey provides observational associations that can reflect confounding, selection bias, and measurement error. Because the SDOH variables are interdependent and the data are observational, the quantities needed to parameterize these mechanisms are best expressed as explicit causal estimands rather than as ad hoc conditional probabilities. The companion paper Hotton et al. (2025) describes the substantive causal structure and mechanism used to represent SDOH effects in INFORM-HIV. In this paper, we focus on the estimation step. In the following paragraph, we describe how we use the *g*-formula (Naimi et al. 2017) to estimate the required effect quantities from survey data, quantify uncertainty, and translate those estimates into simulator inputs.

### G-computation for counterfactual SDOH effects

The survey data are used to initialize baseline SDOH states for agents in the synthetic population. The variables considered at baseline are employment status (*E*), housing instability (*H*), depression (*D*), and substance use (*S*), along with baseline covariates *L* (age). The goal is to generate agent-level SDOH states that reproduce both the marginal prevalences and the dependence structure observed in the survey data. Because the simulator requires operational conditional probabilities rather than observational associations, we construct these probabilities using the g-formula (Naimi et al. 2017; Mooney et al. 2021). Under an assumed causal ordering as shown in Figure 3 the joint distribution of SDOH states can be factorized as

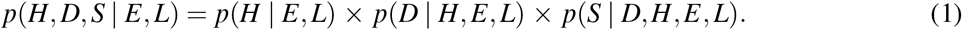

**Figure 3:**
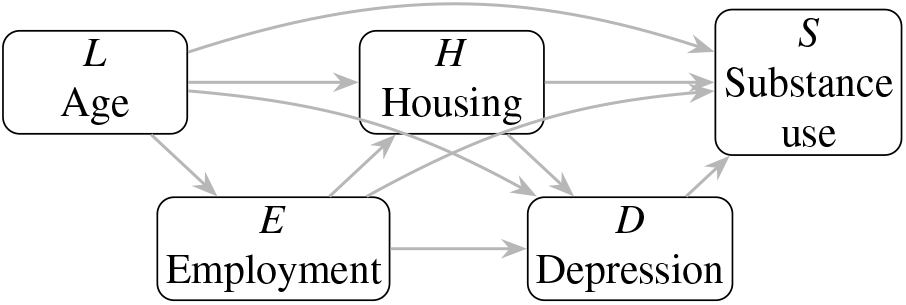
Causal diagram used for baseline SDOH initialization. Age (*L*) is treated as a baseline covariate, and employment (*E*), housing instability (*H*), depression (*D*), and substance use (*S*) are generated sequentially according to the assumed causal ordering.

Each factor is estimated from the survey data using logistic regression models corresponding to the causal parents of each variable. For example, housing instability is modeled as

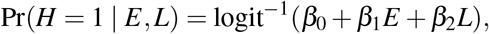

with analogous models for depression and substance use including the appropriate upstream SDOH variables.

Baseline SDOH states are then assigned sequentially for each agent. Given agent attributes (*E*_*i*_, *L*_*i*_), housing is first sampled from the estimated probability Pr(*H*_*i*_ = 1 |*E*_*i*_, *L*_*i*_). Depression is then sampled conditional on (*H*_*i*_, *E*_*i*_, *L*_*i*_), followed by substance use conditional on (*D*_*i*_, *H*_*i*_, *E*_*i*_, *L*_*i*_). In effect, each agent’s baseline SDOH configuration is drawn from the estimated distribution 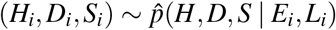.

### 3.2 Dimension Reduction via Morris Screening

We initially considered a broad parameter space comprising 21 uncertain inputs for the INFORM-HIV model. Performing calibration in the full parameter space would be computationally demanding and could lead to weak identifiability among parameters (Semochkina and Walsh 2025). To address this issue, we conducted Morris screening (Morris 1991), a global sensitivity screening step prior to calibration to identify the most influential inputs and reduce the effective dimension of the inference problem. The method evaluates the influence of each input parameter by computing *elementary effects*, which measure changes in the model output induced by one-at-a-time perturbations of parameters across the input space.

The Morris design constructs random trajectories in the parameter space, perturbing one parameter at a time while holding others fixed. Repeating this yields a distribution of elementary effects for each parameter. We summarize these using the mean absolute effect, 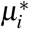 which reflects overall influence, and the standard deviation, *σ*_*i*_, which indicates nonlinearity or interactions. We used *r* = 20 trajectories and *p* = 6 grid levels, requiring *r*×(*k* + 1) simulator evaluations. Sensitivity was assessed with respect to the average HIV prevalence over the last 5 years. Based on joint inspection of 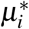 and *σ*_*i*_ (Figure 4), parameters with consistently low 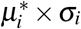 were excluded from the subsequent calibration, reducing the parameter space from 21 to five key inputs *θ*_1_, *θ*_5_, *θ*_6_, *θ*_12_, *θ*_13_, corresponding to probability of transmission for insertive partner, weekly frequency of sex with casual partners, weekly frequency of sex with main partners, PrEP use prevalence, and probability of transmission for receptive partner respectively. These parameters define the calibration space for the subsequent Bayesian inference procedure.

**Figure 4:**
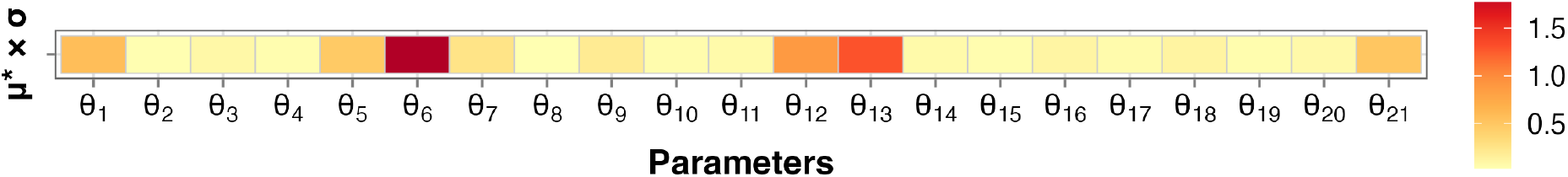
Heatmap of composite Morris sensitivity indices (*μ*^*\**^× *σ*) for 21 model parameters for average HIV prevalence at 5 years as the quantity of interest.

### 3.3 Bayesian Calibration

After input dimension reduction, we calibrated the model against observed epidemiological data. The goal of calibration is to identify parameter combinations that reproduce the current HIV burden in the target population. Rather than seeking a single optimal parameter vector, we characterize the set of parameter values that are consistent with the observed prevalence while accounting for input uncertainty in the simulator (Kennedy and O’Hagan 2001; Andrianakis et al. 2017). Let *θ*∈Θ ⊂ℝ^*d*^ denote the reduced parameter vector with *d* = 5. Let *f*_*t*_(*θ, ω*) denote the simulated HIV prevalence at time *t* under parameter *θ* and stochastic seed *ω*. The simulator is stochastic, so repeated runs at the same parameter value produce distinct trajectories. We denote the simulator as *f* : Θ × Ω *→*ℝ^*T*^, where *T* is the number of simulated time points (years). In this specific application *T* was 20.

The calibration target was the observed current HIV prevalence, defined as the empirical average prevalence over the last five years, *y*_obs_ = 0.294. For a fixed parameter *θ*, we generate *R* independent stochastic trajectories and compute the time-averaged prevalence over the last five simulated years for each replicate, 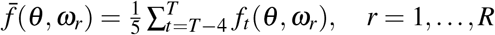. The mean response at each unique *θ* is then obtained by 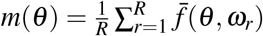.

The calibration task can therefore be viewed as an inverse problem in which the unknown parameter vector *θ* is inferred from the observed prevalence *y*_obs_. Because the mapping *θ* ↦ *m*(*θ*) is nonlinear and generally non-injective, multiple parameter configurations may produce similar prevalence values. A deterministic formulation would identify a single solution that minimizes the discrepancy between simulated and observed prevalence, but such a point estimate does not capture the range of parameter values that are consistent with the data. Instead, in a Bayesian formulation, a prior distribution *π*(*θ*) is specified over the parameter space and the calibration target is modeled as *y*_obs_ = *m*(*θ*) + *ε*, where *ε* represents observation or model error (Baker et al. 2022). The resulting posterior distribution *π*(*θ*| *y*_obs_) characterizes the set of parameter configurations that reproduce the observed epidemic level while accounting for uncertainty.

In practice, traditional posterior inference (e.g., SMC, MCMC) can be computationally expensive (Speich et al. 2021). To address this challenge, we adopt a simulation-based inference (SBI) framework (Dyer et al. 2024) in which the simulator is treated as an implicit generative model linking parameters to observable summaries through forward simulation. In this setting, parameter values are drawn from a prior distribution and the simulator is used to generate the corresponding outputs. The resulting simulated parameter–output pairs form a training dataset for neural posterior estimation (NPE), which learns a flexible approximation to the posterior distribution of the parameters conditional on the observed summary statistic. In our application, the summary statistic is the simulated mean prevalence *m*(*θ*). Training data consist of pairs (*θ*_*i*_, *m*(*θ*_*i*_)) generated from the simulator, and a neural conditional density estimator is trained to approximate *π*(*θ*|*m*(*θ*)). Sequential NPE refines the sampling distribution toward regions of the parameter space that produce prevalence values close to the observed target of 0.294, yielding an approximate posterior distribution that characterizes the set of parameter configurations consistent with the observed epidemic state.

#### Simulation Campaign

To generate training data for calibration and posterior approximation, we conducted a simulation campaign over the reduced five-dimensional parameter space Θ ⊂ ℝ^5^ defined by the Morris screening. We constructed a Latin hypercube design (Sacks et al. 1989) of size *N* = 100 over Θ to obtain space-filling coverage of the parameter space. At each design point *θ*_*i*_, *i* = 1, …, 100, the INFORM-HIV model was executed *R* = 10 times using independent random seeds, producing a total of 1000 simulated trajectories. The same set of seeds was repeated across parameter settings to control stochastic variability. For each run we recorded annual HIV prevalence and computed the summary statistic used for calibration, namely the mean prevalence over the final five simulated years. We summarized each parameter setting by the mean trajectory *m*(*θ*). The resulting dataset of parameter–summary pairs served as the training data for the SBI procedure.

#### Calibration Results

The posterior distribution obtained via sequential neural posterior estimation characterizes parameter configurations that reproduce the observed five-year average HIV prevalence of 29.4%. The left panel of Figure 5 shows the 1D and 2D posterior distributions of the five parameters. The marginal densities on the diagonal exhibit multimodal behaviors for at least two parameters, and the pairwise plots reveal nonlinear dependence patterns, and multiple high-density regions. We further sampled 100 parameter vectors from the posterior and performed forward simulations with *R* = 10 stochastic replicates at each setting. The resulting prevalence trajectories are shown in the middle panel of Figure 5, and the histogram on the right panel summarizes the posterior distribution over the calibrated mean HIV prevalence.

**Figure 5:**
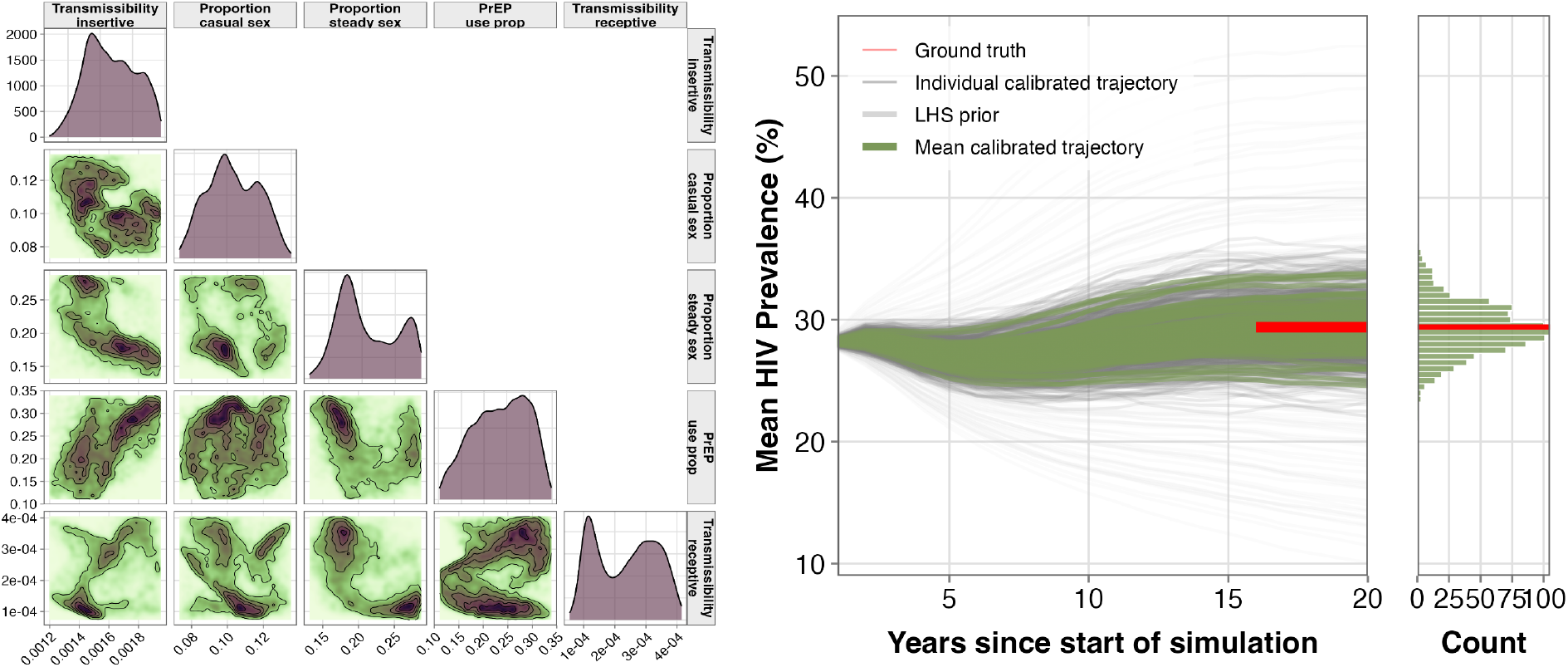
Posterior parameter distributions and calibrated HIV prevalence trajectories. Left: pairwise posterior of five parameters (marginals on the diagonal, 2D KDE contours off-diagonal). Center: prior trajectories (light grey), individual calibrated runs (thin colored), and posterior mean trajectories (bold). Right: marginal distribution of prevalence at year 20 across calibrated runs, with the red line indicating the target.

**Figure 6:**
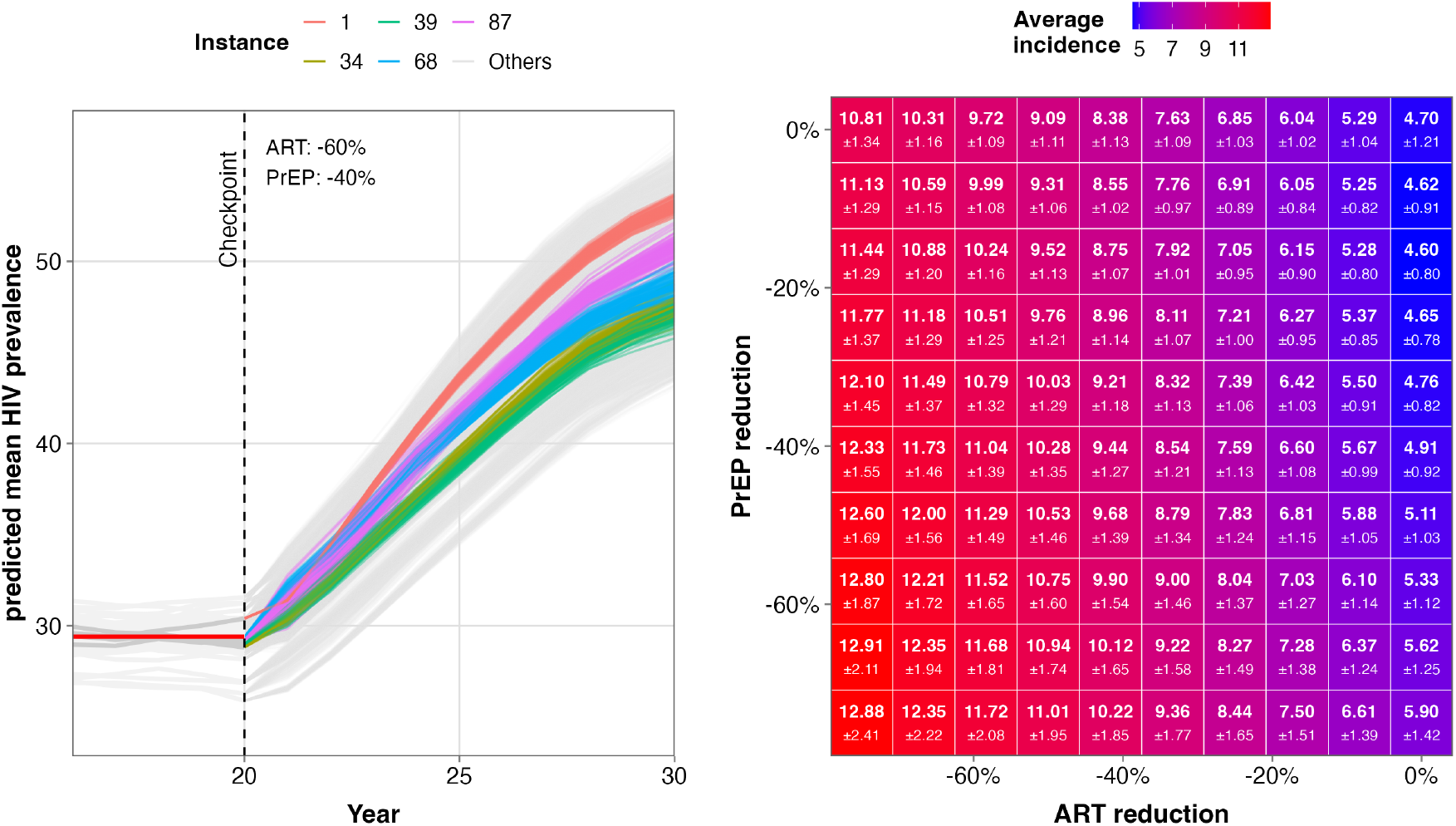
Surrogate-based projections of HIV prevalence and incidence under ART and PrEP reduction scenarios. Left: Prevalence trajectories under a 60% ART and 40% PrEP reduction from five representative posterior checkpoints. Grey lines before the checkpoint (year 20) show calibrated histories; after the checkpoint, colored lines denote the five shown instances and grey lines denote the remaining posterior projections. The red segment marks the 29.4% prevalence at the checkpoint. The predictive distribution comprises all post-checkpoint trajectories. Right: Heatmap of mean HIV incidence (per 100 person-years) over a 10-year horizon across ART and PrEP reductions, with each cell reporting the mean (standard deviation) over posterior samples and surrogate uncertainty.

### 3.4 ART and PrEP use Scenario Exploration

Once the model is calibrated, it can be used to address policy-relevant questions by evaluating the effects of changing policy levers in the model. In our application, a key question is how disruptions in biomedical prevention and care, specifically reductions in ART and PrEP use, may influence future HIV outcomes. While such questions could be explored by simulating the calibrated model forward in time from a single calibrated state under alternative intervention settings, it is important to account for uncertainty in the calibrated parameters. The posterior distribution obtained during calibration represents a set of parameter configurations that are consistent with the observed epidemic state, and therefore forward projections should incorporate this posterior uncertainty rather than relying on a single snapshot of the model state.

To propagate this uncertainty, we conducted forward simulations from calibrated model states. Specifically, we retained 100 posterior trajectories as checkpointed system states at the calibration horizon (year 20). Each checkpoint encodes the epidemiological state of the system at the end of the historical period. These checkpointed states were then used as initial conditions for forward simulations into future years.

We consider counterfactual scenarios defined by proportional reductions in ART coverage and PrEP use. For a given checkpointed state and a specified pair of reduction levels (*r*_ART_, *r*_PrEP_), the model can be propagated forward to obtain projected prevalence and incidence trajectories over the next 10 years. Repeating this procedure across the 100 checkpointed states yields a distribution of outcomes that reflects posterior uncertainty in model parameters and latent states.

However, direct simulation across a fine grid of intervention combinations is computationally prohibitive. To characterize the joint impact of varying ART and PrEP reductions over a continuous two-dimensional intervention space, one would need to repeat forward simulations for many combinations of (*r*_ART_, *r*_PrEP_) and for each checkpointed model state. The resulting computational cost scales linearly with the number of intervention scenarios and posterior samples, making comprehensive policy exploration impractical without additional approximation methods.

#### 3.4.1 Multi-surrogate assisted intervention scenario exploration

To reduce the computational cost of evaluating intervention scenarios, we construct surrogate models (Gramacy 2020) and emulate the mapping from intervention inputs (*r*_ART_, *r*_PrEP_) to epidemiological outcomes. For each checkpointed model state, this mapping to 10-year prevalence and incidence trajectories is approximated using Gaussian process (GP) regression (Rasmussen and Williams 2006), which provides both a flexible nonparametric fit and a predictive distribution for uncertainty quantification. Training data are generated via an initial LHS design (Sacks et al. 1989) of 20 intervention combinations with 10 stochastic replicates per combination. Separate GP surrogates are trained for prevalence and incidence, and this procedure is repeated across all 100 checkpointed posterior trajectories, yielding an ensemble of surrogates conditional on calibrated model states. For any intervention, predictions from these surrogates are combined by marginalizing over the posterior ensemble, integrating uncertainty from both calibration and surrogate approximation. We refer to this ensembling of surrogates as the multi-surrogate approach.

##### Construction of Individual Gaussian Process surrogates for Time-Series Output

Let **u** = (*r*_ART_, *r*_PrEP_) ∈ 𝒰 ⊂ ℝ^2^ denote the intervention input and let *y*(**u**) represent an outcome such as HIV prevalence. A GP surrogate models the response as *y*(**u**) = *μ*(**u**) + *Z*(**u**), where *Z*(**u**) is a zero-mean GP with covariance kernel *k*(**u, u**′). Given training data 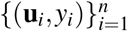, the GP provides a predictive Gaussian distribution at new inputs, with hyperparameters estimated via marginal likelihood. In our application the simulator output is a time series **y**(**u**) ∈ ℝ^*T*^ with *T* = 10. Rather than modeling each time point independently, which inflates the size of the regression problem and increases computational cost, we represent trajectories using a low-dimensional basis decomposition

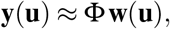

where Φ = [*φ*_1_, …, *φ*_*J*_] ∈ ℝ^*T*×*J*^ contains basis vectors obtained from singular value decomposition of simulated trajectories and **w**(**u**) denotes the corresponding weights (Fadikar et al. 2018). Independent GP models are then fitted to each weight function *w*_*j*_(**u**). Predictions for a new intervention input **u**_*\**_ are obtained by predicting the weights using the GP models and reconstructing the trajectory as **ŷ**(**u**_*\**_) = Φ **ŵ** (**u**_*\**_), with uncertainty propagated through the basis representation.

##### Accounting for Posterior Uncertainty in the Multi-Surrogate setup

Let *θ* denote the simulation parameter vector with posterior distribution *π*(*θ* | *y*_obs_). For an intervention input **u**, the GP surrogate trained at a checkpointed model state, which corresponds to a posterior sample *θ*^*\**^ from *π*(*θ* | *y*_obs_) provides an approximate predictive distribution *p*_GP_(**y**(**u**) |*θ*^*\**^). Thus, the overall predictive distribution accounting for calibration uncertainty in this multi-surrogate approach is obtained by marginalizing over the posterior,

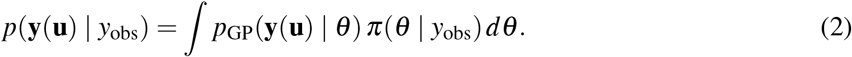

In practice this integral is approximated using Monte Carlo sampling. Let 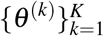 denote posterior samples retained from calibration. For each *θ*^(*k*)^, the corresponding GP surrogate produces a Gaussian predictive distribution for the outcome trajectory. Samples drawn from these predictive distributions are aggregated across posterior samples to approximate *p*(**y**(**u**)|*y*_obs_). Posterior predictive summaries, such as mean trajectories and credible intervals, are then computed from the pooled samples, thereby incorporating both calibration uncertainty and surrogate approximation uncertainty.

This marginalization enables the propagation of multiple sources of uncertainty into the projected outcomes. In stochastic simulators such as INFORM-HIV, uncertainty arises from epistemic sources, including uncertain model parameters, noise in survey data, calibration targets, and surrogate approximation error, as well as from intrinsic stochastic variability in simulator dynamics. These components are represented probabilistically and combined through the marginalization to obtain predictive distributions for future outcomes (eq. (2)). This provides a practical and scalable approach for generating uncertainty-aware projections while retaining the full structure of the underlying uncertainties.

#### 3.4.2 Results: Impact of ART and PrEP reduction on HIV incidence

In the PrEP and ART funding reduction scenarios, ART and PrEP were decreased relative to baseline (calibrated) levels over 1 year and the reduced levels were maintained over the following 9 years. Final outcomes are presented based on data from the 10th year of the simulation post-checkpoint. HIV incidence rates for each scenario were computed by summing the total number of new infections over annual intervals and dividing by the total person-time contribution among at-risk agents during that period. Under a baseline scenario of no change in funding, predicted HIV incidence at 10 years was 4.60 (standard error; se = 0.62) per 100 person-years. A 20% reduction in ART with no reduction in PrEP resulted in an increase in incidence to 6.04 (se = 0.52) per 100 person years, whereas a 20% reduction in both ART and PrEP yielded incidence of 6.14 (se = 0.46) per 100 person-years. A 40% reduction in ART and PrEP resulted in a doubling of HIV incidence to 9.44 (se = 0.65) per 100 person-years over 10 years.

These results demonstrate that reductions in HIV program funding could substantially increase HIV transmission. The impact of PrEP reductions were less pronounced when ART levels were maintained but became more important in scenarios with larger decreases in ART. This is not surprising, as the impact of PrEP is expected to be greater in settings with higher incidence or lower levels of viral suppression. Of note, this is a simple example provided for demonstration purposes. The substantive findings should be interpreted with caution since they are dependent on assumptions about ART and PrEP adherence, baseline population coverage levels, and whether PrEP is distributed randomly or according to individual or partner characteristics. In this example, PrEP was distributed randomly and was not preferentially given to those at elevated risk, so its impact is likely underestimated.

## 4 DISCUSSION

This work presents the DDUADS workflow for decision support using complex stochastic epidemic models. Using the INFORM-HIV model as a case study, we demonstrated how heterogeneous observational data, uncertain model inputs, and computationally intensive simulations can be integrated into a coherent pipeline for flexible and uncertainty-aware scenario analysis. The workflow combines causal translation of survey data into simulator mechanisms, global sensitivity screening for dimension reduction, Bayesian calibration using simulation-based inference, and surrogate-assisted exploration of intervention scenarios. This application has important implications for modeling HIV and other infectious diseases that occur within complex social systems, where policy makers are increasingly focused on interventions to address social determinants of health in addition to biomedical interventions. Simulating these interventions requires significant increases in mechanistic complexity and uncertainty compared to prior approaches. The workflow presented here facilitates more systematic and thorough scenario exploration for complex ABMs.

We re-emphasize the role of uncertainty propagation during scenario exploration. Intervention projections are evaluated across an ensemble of parameter samples drawn from the posterior distribution obtained during calibration. Since each surrogate prediction is Gaussian, the resulting predictive distribution can be interpreted as a mixture of Gaussian components induced by the posterior ensemble. This representation can approximate complex output distributions, including skewed or multimodal patterns that arise in nonlinear systems, without imposing restrictive assumptions on the form of the output distribution. As a result, the framework seeks to provide a more faithful characterization of uncertainty in projected epidemic trajectories under alternative interventions.

There are several directions for further development. Calibration in this work is based on a low-dimensional summary of the data, which may not fully encapsulate all important model dynamics. In particular, instead of using the mean trajectory (or other summary statistics) from replicates as the simulator output in the calibration routine, trajectory-oriented approaches (Fadikar et al. 2023) may provide more efficient and better quantification of uncertainties by identifying parameter and random stream combinations that reproduce observed dynamics. In addition, the surrogate models are constructed independently for each checkpointed state, which may limit scalability as the number of posterior samples increases. Extensions based on shared or hierarchical surrogate models could improve efficiency in such settings.

## Data Availability

All data produced in the present study are available upon reasonable request to the authors

## ACKNOWLEDGMENTS

This research was funded by the National Institutes of Health/National Institute of Drug Abuse under grant number R01DA057350. This material is based upon work supported by the U.S. Department of Energy, Office of Science, under contract number DE-AC02-06CH1135). This work was completed in part with resources provided by the University of Chicago’s Research Computing Center.

